# Performance of the Verily Study Watch for Measuring Sleep Compared to Polysomnography

**DOI:** 10.1101/2024.09.10.24313427

**Authors:** Sohrab Saeb, Benjamin W. Nelson, Poulami Barman, Nishant Verma, Hannah Allen, Massimiliano de Zambotti, Fiona C. Baker, Nicole Arra, Niranjan Sridhar, Shannon S. Sullivan, Scooter Plowman, Erin Rainaldi, Ritu Kapur, Sooyoon Shin

## Abstract

**Introduction:** This study evaluated the performance of a wrist-worn wearable, Verily Study Watch (VSW), in detecting key sleep measures against polysomnography (PSG).

**Methods:** We collected data from 41 adults without obstructive sleep apnea or insomnia during a single overnight laboratory visit. We evaluated epoch-by-epoch performance for sleep versus wake classification, sleep stage classification and duration, total sleep time (TST), wake after sleep onset (WASO), sleep onset latency (SOL), sleep efficiency (SE), and number of awakenings (NAWK). Performance metrics included sensitivity, specificity, Cohen’s kappa, and Bland-Altman analyses.

**Results:** Sensitivity and specificity (95% CIs) of sleep versus wake classification were 0.97 (0.96, 0.98) and 0.70 (0.66, 0.74), respectively. Cohen’s kappa (95% CI) for 4-class stage detection was 0.64 (0.18, 0.82). Most VSW sleep measures had proportional bias. The mean bias values (95% CI) were 14.0 minutes (5.55, 23.20) for TST, - 13.1 minutes (-21.33, -6.21) for WASO, 2.97% (1.25, 4.84) for SE, -1.34 minutes (-7.29, 4.81) for SOL, 1.91 minutes (-8.28, 11.98) for *light sleep* duration, 5.24 minutes (-3.35, 14.13) for *deep sleep* duration, and 6.39 minutes (-0.68, 13.18) for *REM sleep* duration. Mean and median NAWK count differences (95% CI) were 0.05 (-0.42, 0.53) and 0.0 (0.0, 0.0), respectively.

**Discussion:** Results support applying the VSW to track overnight sleep measures in free-living settings. Registered at clinicaltrials.gov (NCT05276362).

## Introduction

Characterizing sleep in a free-living setting provides valuable insights into physical and mental health. Changes in sleep may be key in the diagnosis of sleep disorders like insomnia and hypersomnia, and are clinically meaningful components for tracking mental and cardiovascular health, as well as other conditions (Parish, 2009)(Freeman et al., 2020)(Tobaldini et al., 2019)(Young et al., 2008)(Ahmadi et al., 2009)(Hayashino et al., 2010). The gold standard for sleep assessment is lab-based polysomnography (PSG). However, PSG is resource intensive, challenging to administer and subject to intra- and inter-scorer variability, moreover, availability of PSG laboratories may be limited (Norman et al., 2000)(Deutsch et al., 2006). It is also impractical for long-term surveillance, and may be prone to artifacts that affect representativeness, such as altered sleep patterns due to the novelty of a laboratory, and/or the discomfort of the electrode setting (Toussaint et al., 1995). Furthermore, while portable PSG tools do exist, they still have limited application in free-living environments or routine clinical care.

Wearable sensors, particularly wrist-worn devices, provide a promising avenue for sleep assessment in free-living settings. These devices are widely available, relatively inexpensive, comfortable to wear during sleep and include physiological sensors, such as photoplethysmogram (PPG) and accelerometer, that can be used for sleep monitoring (Imtiaz, 2021)(de Zambotti et al., 2024). However, before utilizing wearable-based technology as a routine approach to monitor daily sleep, whether for care or for research purposes, it is important to conduct performance evaluation of devices and algorithms compared to a gold standard reference such as PSG. Furthermore, researchers now know the importance of conducting those analytical and clinical evaluations across diverse and representative populations, such as participants with different ages or skin tones, to increase confidence in the generalizability of the results (Colvonen et al., 2020)(Baumert et al., 2023)(Nelson et al., 2020).

This study evaluated the performance of the Verily Study Watch (VSW, a wrist-worn wearable) to monitor sleep in a diverse cohort of sleepers without obstructive sleep apnea (OSA) or elevated insomnia symptoms, by comparing VSW sleep measures against measures obtained from PSG-based labels. The VSW classifies every 30-second epoch into 4 sleep-related stages: wake, light sleep, deep sleep, and rapid eye movement (REM) sleep. These classifications enable the calculation of multiple sleep measures that provide information on the quantity and the quality of an individual’s overnight sleep. In this study, the measures of interest were: total sleep time (TST), wake after sleep onset (WASO), sleep efficiency (SE), sleep onset latency (SOL), number of awakenings (NAWK), and duration of each sleep stage. Our main objective was to compare epoch-by-epoch VSW-against PSG-derived classification of sleep-versus-wake state and of sleep stages. Additionally, we wanted to assess the VSW’s accuracy for all computed sleep measures (listed above). Finally, we wanted to evaluate any potential variability in the performance of the VSW’s sleep algorithm across demographic factors such as age, sex, body mass index (BMI), skin tone, and arm hair density.

## Methods

### Participants

The basic setup and eligibility for the study have been described elsewhere (Nelson, 2024, submitted). Eligible participants were between 18-80 years old, agreed to abstain from any drugs or medications that may affect sleep or wakefulness prior to and during the lab visit, and did not have identified symptoms of sleep disorders, such as obstructive sleep apnea (OSA, defined by OSA 50 score ≥5), or elevated insomnia symptoms (defined by having an insomnia severity index (ISI) score ≥ 8). The study was approved by the WCG Institutional Review Board (20215892), and all participants provided informed consent. This study was registered at clinicaltrials.gov (NCT05276362).

### Data Collection

For each participant, data were collected during a single overnight stay in a sleep laboratory at a single site (SRI; Menlo Park, California), between February 14th and September 1st, 2023. Participants slept in comfortable, sound-proof and temperature-controlled bedrooms. Standard PSG protocols were used for preparation, recording procedures, and instrument calibration (Nelson, 2024, submitted).

### Study Watch Data

During their overnight visit, participants wore the VSW on their dominant wrist. This analysis was part of a larger study including two devices: the Verily Numetric Watch (VNW) (Nelson, 2024, submitted), in addition to the VSW. VSW is equipped with two sensors: a green-light PPG sensor, and a 3-axis accelerometer. Both sensors had a sampling frequency of 60 Hz (in the VNW, the PPG sensor consists of a green light emitter diode and two PPG signal channels and the sampling rate of the 3-axis accelerometer is 104 Hz). Using the PPG and accelerometer signals, the VSW classifies every 30-second epoch into one of the following 4 classes: *wake, light sleep, deep sleep*, and *REM sleep*.

The sleep stage classification algorithm consisted of a deep convolutional neural network that was initially trained using 10,000 nights of data from the Sleep Heart Health Study (SHHS) and Multi-Ethnic Study of Atherosclerosis (MESA) public datasets (Sridhar et al., 2020). The algorithm was fine-tuned using a smaller dataset collected at SRI, consisting of 30 nights of PSG-labeled data.

The overnight sleep measures, including TST, WASO, SE, SOL, NAWK, and sleep stage durations (Supplementary Table 1), for each participant were calculated using the VSW’s predicted sleep stages, from the time the lights were turned off (“lights-off”) to the time lights were turned back on (“lights-on”). VSW start time was synced to the Lights Off time recorded on PSG to ensure alignment for analysis of simultaneously recorded signals, using procedures described elsewhere (Nelson, 2024, submitted) (de Zambotti et al., 2019).

### Reference Data

Standard laboratory PSG sleep assessment including electroencephalography (EEG), submental electromyography and bilateral electrooculography was performed according to the American Academy of Sleep Medicine (AASM) guidelines. Leg movement, electrocardiography (ECG), respiratory, and oxygen saturation signals were also collected and used to confirm the absence of sleep disordered breathing. All recordings were performed using the Compumedics Grael^®^ HD-PSG system (Compumedics, Abbotsford, Victoria, Australia). Two independent sleep scorers labeled every 30-second epoch of the PSG data by one of the following categories: *wake, N1, N2, N3, REM*. Inter-rater reliability (Kappa) between the two scorers was 91%, and discrepancies were resolved by a third scorer.

For this analysis, PSG stages *N1* and *N2* were combined into a single *light sleep* category, and PSG *N3* was termed *deep sleep*.

Similar to VSW, for each participant, the overnight sleep measures for PSG were calculated using the sleep scorer’s stage labels from lights-off to lights-on.

### Performance Evaluation

Performance evaluation was done based on an existing standardization framework (Menghini et al., 2021).

We evaluated the epoch-by-epoch performance of VSW’s sleep stage classification against PSG in two ways: (1) *sleep* versus *wake* classification, using *sleep* as the positive class; and (2) 4-class (wake, light, deep, REM) sleep stage classification. For the evaluation of sleep vs wake classification, we estimated sensitivity, specificity, positive predictive value (PPV), and negative predictive value (NPV). We calculated the 95% CI using cluster bootstrapping, and we accounted for the clustering of epochs within a participant using logistic mixed-effect regression models with the participant as random effect. For the 4-class stage classification, we used Cohen’s kappa and accuracy along with their 95% bootstrapped CIs. Additionally we evaluated performance for each sleep stage by reporting Cohen’s Kappa, accuracy, PPV and sensitivity using the average method (Menghini et al., 2021). To obtain performance metrics on each sleep stage, the outcomes were dichotomized to the sleep stage of interest against all others. The average method calculates kappa for each individual participant and then averages out the kappa across all participants with their associated bootstrapped 95% CIs. All analyses were confined to the lights-off to lights-on period.

For evaluating the performance of all overnight sleep measures except NAWK, we performed the Bland Altman analysis, estimating the mean bias and lower and upper limits of agreement, testing for the assumptions of proportional bias, heteroscedasticity, and normality. For NAWK, we estimated the mean and median count difference and linearly weighted Cohen’s kappa with their 95% CIs.

Finally, we evaluated all performance metrics across the participant subgroups, including age, sex, BMI, skin tone, arm hair index. For subgroups with insufficient number of samples (< 10), we did not evaluate the performance.

All analyses were performed with R version 4.3.1 (2023-06-16).

## Results

There were 41 adult participants (18 male, age range: 18-78 years) in this study. Participants had a diverse range of skin tones, BMI, and arm hair density (Supplementary Table 2).

VSW estimated sleep stages for a total of 38,796 epochs with data collected between lights-off and lights-on for each participant.

The sensitivity (95% CI) of the VSW in classifying sleep vs wake was 0.97 (0.96, 0.98), specificity (95% CI) was 0.70 (0.66, 0.74), PPV (95% CI) was 0.93 (0.92, 0.95), and NPV (95% CI) was 0.83 (0.78, 0.88) (Table 1).

**Table 1.** Performance of VSW’s sleep vs wake classification against PSG reference.

|  | Sensitivity (95% CI) | Specificity (95% CI) | NPV (95% CI) | PPV (95% CI) |
| --- | --- | --- | --- | --- |
| **Sleep vs Wake** | 0.97 (0.96, 0.98) | 0.70 (0.66, 0.74) | 0.83 (0.78, 0.88) | 0.93 (0.92, 0.95) |

CI: Confidence Interval; NPV: Negative Predictive Value; PPV: Positive Predictive Value

The accuracy (95% CI) of the VSW sleep algorithm in classifying all 4 sleep stages was 0.78 (0.58, 0.89), and the kappa (95% CI) was 0.64 (0.18, 0.82) (Table 2). There was variability in the performance across different sleep stages, with *light sleep* stage prediction having the lowest accuracy (Table 2), as there were instances of confusion between the *light sleep* stage and all other stages (Supplementary Table 3).

**Table 2.** VSW’s performance in 4-class sleep stage detection against the PSG reference.

| Sleep Stage | Kappa (95% CI) | Accuracy (95% CI) | PPV (95% CI) | Sensitivity (95% CI) |
| --- | --- | --- | --- | --- |
| Overall | 0.64 (0.18, 0.82) | 0.78 (0.58, 0.89) | NA | NA |
| Wake | 0.70 (0.43, 0.90) | 0.92 (0.76, 0.98) | 0.82 (0.51, 0.98) | 0.71 (0.45, 0.94) |
| Light | 0.60 (0.29, 0.78) | 0.80 (0.66, 0.89) | 0.80 (0.55, 0.91) | 0.81 (0.59, 0.94) |
| Deep | 0.66 (0.17, 0.91) | 0.92 (0.84, 0.98) | 0.69 (0.09, 0.97) | 0.77 (0.37, 0.98) |
| REM | 0.74 (0.38, 0.90) | 0.92 (0.82, 0.98) | 0.76 (0.44, 0.96) | 0.84 (0.47, 0.99) |

CI: confidence interval; PPV: Positive Predictive Value; REM: Rapid Eye Movement

**Table 3.**
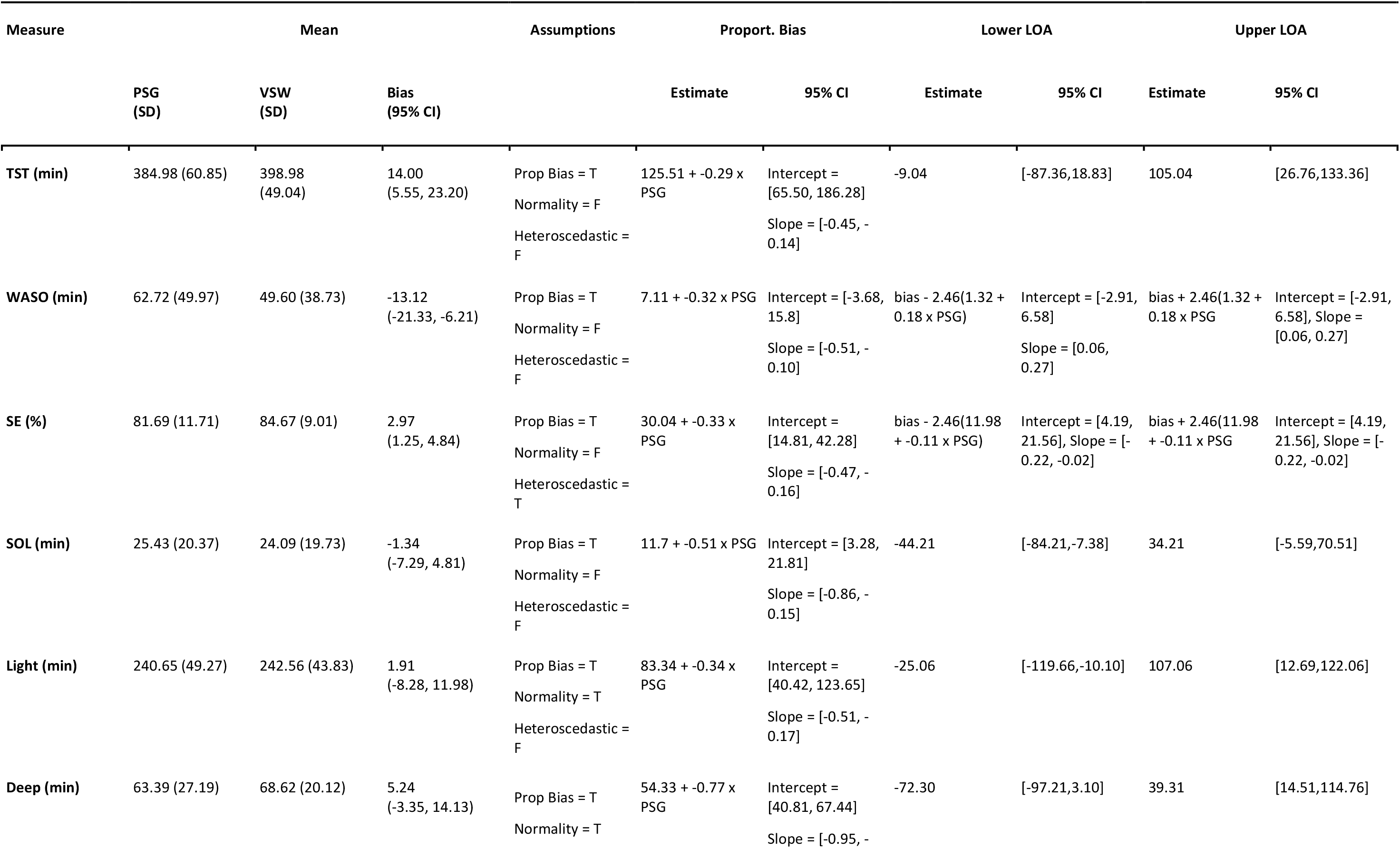
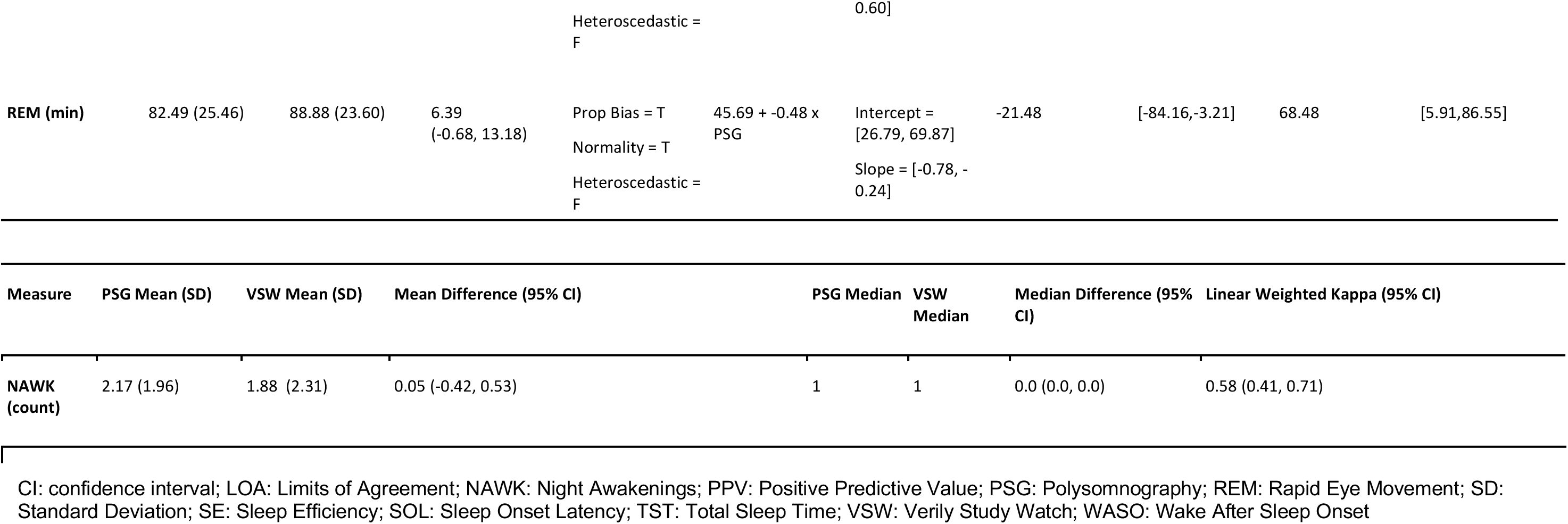
Performance of VSW overnight sleep measures against PSG reference.

Mean bias and 95% CI values for all overnight sleep measures is shown in Table 3. Bland-Altman analyses (Figure 1) showed that all measures had significant proportional bias, with the VSW overestimating the measures at the lower end of the distribution, and underestimating them at the upper end, relative to the PSG. For all overnight sleep measures except the sleep stage durations, the assumption of normality was false, and for all measures except SE the assumption of homoscedasticity was true.

**Figure 1.**
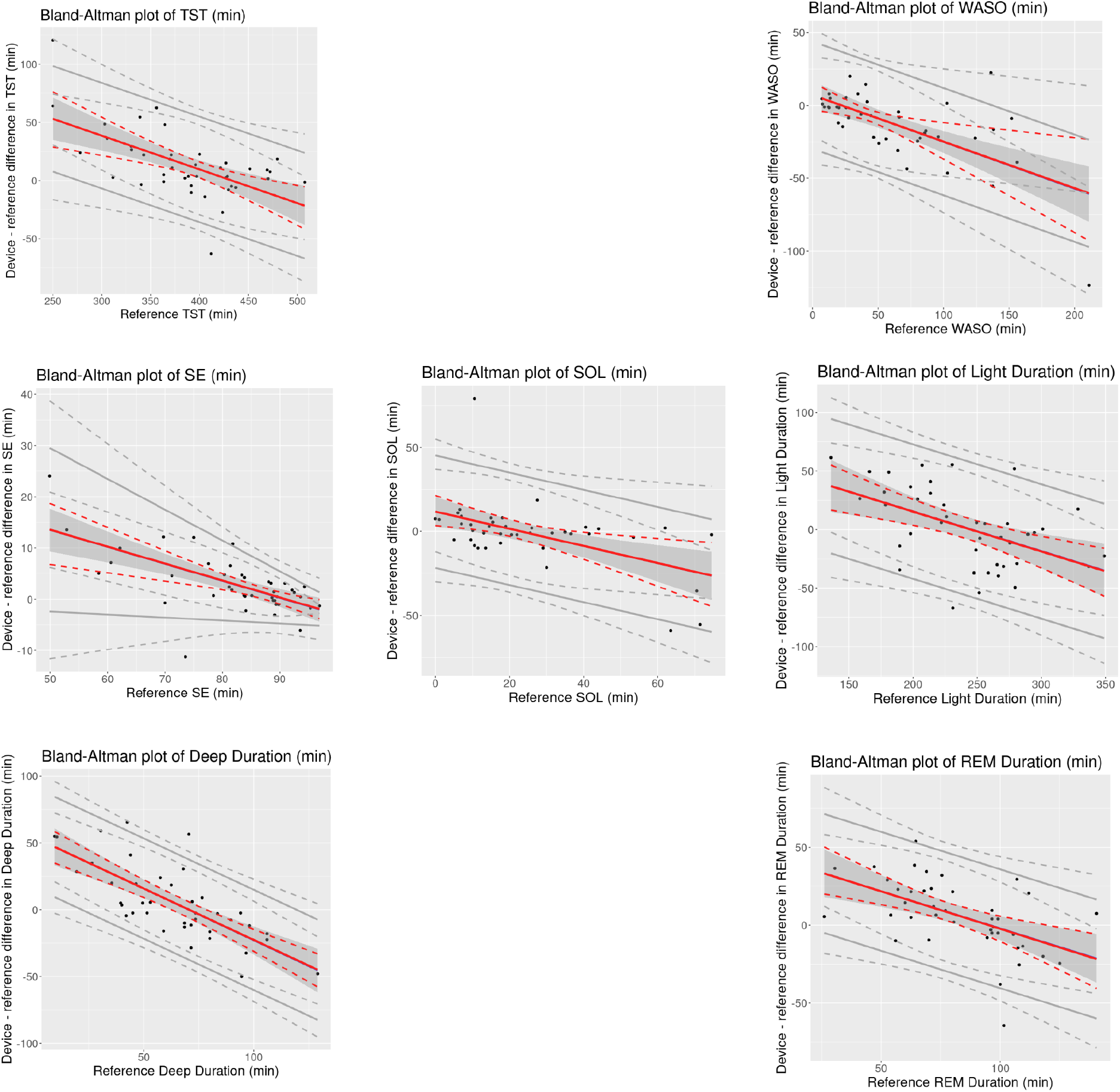
Bland-Altman plots of overnight sleep measures for the device (VSW) against the reference (PSG). Solid red lines indicate mean bias, dotted red lines indicate 95% CI of mean bias, solid gray lines indicate the 95% LOAs, and dotted gray lines indicate 95% CI of LOAs. Black dots are observations. (CI: confidence interval; REM: Rapid Eye Movement; SD: Standard Deviation; SE: Sleep Efficiency; SOL: Sleep Onset Latency; TST: Total Sleep Time; WASO: Wake After Sleep Onset)

Performance of the VSW metrics across demographic subgroups of age, sex, BMI, skin tone, and arm hair density are reported (Supplementary Tables 4 and 5) without formal statistical testing, due to small subgroup sample size.

## Discussion

The results of this study show the ability of the VSW to capture information related to sleep quantity and quality, as well as the distribution of sleep stages across overnight periods in individuals without OSA or elevated insomnia symptoms. The sensitivity and specificity of the VSW in classifying sleep vs wake were 0.97 and 0.70 respectively, and the Cohen’s kappa for the 4-class stage classification was 0.64. This performance supports the application of the VSW to monitor overnight sleep in free-living settings.

As with other wearable sleep-wake detection devices (Pesonen and Kuula, 2018)(de Zambotti et al., 2016)(Miller et al., 2022), the sleep algorithm in this study was more likely to miss wake than sleep, as reflected in the higher sensitivity relative to specificity, and the positive and negative bias values for TST and WASO, respectively. When evaluating the performance of sleep monitoring devices, the AASM has established a range of ‘allowable differences’, based on actigraphy studies conducted in patients with specific sleep disorders (e.g. insomnia)(Smith et al., 2018). The 95% CIs of the mean bias estimates for TST, WASO, SOL, and SE measured by the VSW were within those allowable difference ranges. However, for the *proportional* mean bias estimates, which account for variations in bias over the range of measurement, 95% CIs exceeded these thresholds at lower and higher ends of the measurements (Figure 1). Nonetheless, applying the AASM standards to these results may require caution. Unlike the studies included in the AASM assessment, the present study excluded (via questionnaire) participants with symptoms of certain sleep disorders.

There are a few caveats to consider when interpreting our results. First, data collection for this study took place at a sleep laboratory, with standardized study boundaries and settings, such as lights-on/off to define the “in bed’ time period when an individual is (in theory) set to sleep. Free-living environments are more organic and complex, and the generation of sleep measures in them may require additional layers of data. Following the prior example, defining “in bed” time may necessitate additional sensor readings, which then would be integrated into the derivation of the measures, particularly sleep stage classification and duration, or SOL.

Another caveat is that participants in this study were free of sleep-related diagnoses and symptoms (such as OSA or heightened insomnia symptoms). Participants with certain clinical conditions may manifest different patterns in their biological signals (e.g., pulse rate) and/or sleep architecture, which could complicate the sleep stage classification task. Future studies should evaluate the performance of VSW in real-world settings and in clinically relevant populations such as individuals with sleep disorders.

In summary, we evaluated the performance of the VSW and its algorithm to classify sleep versus wake state and the four different sleep stages in sleepers without OSA or heightened insomnia symptoms, as well as a series of measures that illustrate the quantity and quality of overnight sleep. The results demonstrate the potential of VSW to classify sleep vs wake states and sleep stages and compute overnight sleep measures when compared to gold-standard PSG measurements. These findings support further application of the VSW to tracking the overnight sleep behaviors in sleepers without OSA or heightened insomnia symptoms in free-living settings.

## Data Availability

Data from this study are not available due to the nature of this program. Participants did not consent for their data to be shared publicly.

## Acknowledgements

Authors wish to acknowledge participants and study personnel that made the study possible.

