## Supplementary Materials for "Performance of the Verily Study Watch for Measuring Sleep Compared to Polysomnography"

Supplementary Table 1. Definitions of overnight sleep measures. These definitions are based on an existing standardization framework (Menghini et al. 2021).

| Metric | Definition |
|---|---|
| **TST** | Total sleep time; total time in minutes classified as sleep (Deep, Light, or REM) between lights-off and lights-on |
| **WASO** | Wake after sleep onset; total time in minutes classified as awake after the first sleep epoch |
| **SE** | Sleep efficiency, defined as percentage of TST over the lights-off to lights-on period |
| **SOL** | Sleep onset latency; total time in minutes classified as wake before the first epoch classified as sleep |
| **NAWK** | Number of awakenings; number of awake periods after the first sleep epoch |
| **Sleep Stage Duration** | Total time in minutes classified as each of the sleep stages (Deep, Light, or REM) between lights-off and lights-on |

NAWK: Night Awakenings; REM: Rapid Eye Movement; SE: Sleep Efficiency; SOL: Sleep Onset Latency; TST: Total Sleep Time; WASO: Wake After Sleep Onset; REM=Rapid Eye Movement

Supplementary Table 2. Participant Characteristics

| | | N=41 |
|---|---|---|
| **Age (years)** | Median (range) | 34.0 (18.0 - 78.0) |
| | Mean (SD) | 40.5 (16.5) |
| **Age categories, n (%)** | 18-40 | 25 (61.0) |
| | 41-80 | 16 (39.02) |
| **Sex, n (%)** | Female | 23 (56.1) |
| | Male | 18 (43.9) |
| **BMI categories, n (%)** | < 25 | 30 (73.2) |
| | ≥ 25 | 11 (26.8) |
| **Skin tone, n (%)** | Light Skin Tone | 21 (51.2) |
| | Medium Skin Tone | 15 (36.6) |
| | Dark Skin Tone | 5 (12.2) |
| **Arm hair index, n (%)** | 1: Little to no visible arm hair, light in color | 17 (41.5) |
| | 2: Visible, fine, arm hair, light to medium color | 16 (39.0) |
| | 3 and 4: Coarse and very coarse arm hair, medium to dark color | 8 (19.51) |
| **Race, n (%)** | American Indian or Alaska Native | 1 (2.4) |
| | Asian | 8 (19.5) |
| | Black of African American | 4 (9.8) |
| | Mixed race | 4 (9.8) |
| | Native Hawaiian or Other Pacific Islander | 1 (2.4) |

| | | |
|---|---|---|
| | Other | 1 (2.4) |
| | White | 22 (53.7) |
| **Ethnicity, n(%)** | Hispanic or Latino | 4 (9.8) |
| | Not Hispanic or Latino | 37 (90.2) |
| **Dominant hand, n (%)** | Ambidextrous | 1 (2.4) |
| | Left | 5 (12.2) |
| | Right | 35 (85.4) |
| **OSA score** | Median (range) | 0.0 (0.0 - 7.0) |
| | Mean (SD) | 1.3 (1.6) |
| **ISI score** | Median (range) | 3.0 (0.0 - 7.0) |
| | Mean (SD) | 3.0 (1.9) |
| **ESS score** | Median (range) | 5.0 (0.0 - 9.0) |
| | Mean (SD) | 5.0 (2.7) |
| **AHI Index** | Median (range) | 1.3 (0.1 - 4.7) |
| | Mean (SD) | 1.7 (1.2) |

AHI=Apnea hypopnea index ; BMI=body mass index; ESS=Epworth sleepiness scale ; ISI=insomnia severity index; OSA=Obstructive Sleep Apnea; SD=standard deviation

Supplementary Table 3. Confusion matrix for the epoch-by-epoch classification of sleep stages.

| | | Device (VSW) | | | | |
|---|---|---|---|---|---|---|
| | | **Wake** | **Light** | **Deep** | **REM** | **Total Reference** |
| **Reference (PSG)** | **Wake** | 5,029 | 869 | 84 | 97 | 6,079 |
| | **Light** | 1,664 | 15,996 | 1,124 | 1,107 | 19,891 |
| | **Deep** | 65 | 1,599 | 3,840 | 34 | 5,538 |
| | **REM** | 470 | 1,269 | 23 | 5,526 | 7,288 |
| **Total Device** | | 7,228 | 19,733 | 5,071 | 6,764 | 38,796 |

PSG: Polysomnography; REM: Rapid Eye Movement; VSW: Verily Study Watch

Supplementary Table 4. Performance of 'sleep vs wake' classification for participant subgroups.

| Subgroup (n) | | n | Sensitivity (95% CI) | Specificity (95% CI) | NPV (95% CI) | PPV (95% CI) |
|---|---|---|---|---|---|---|
| **Age** | **18-40 yrs** | 25 | 0.97 (0.97, 0.98) | 0.74 (0.68, 0.79) | 0.85 (0.80, 0.89) | 0.95 (0.94, 0.96) |
| | **> 40 yrs** | 16 | 0.96 (0.94, 0.98) | 0.65 (0.60, 0.71) | 0.80 (0.71,0.90) | 0.91 (0.88, 0.94) |
| **Sex** | **Female** | 23 | 0.96 (0.95, 0.98) | 0.71 (0.65, 0.76) | 0.78 (0.70, 0.86) | 0.94 (0.92, 0.96) |
| | **Male** | 18 | 0.98 (0.97, 0.98) | 0.68 (0.62, 0.74) | 0.89 (0.84, 0.92) | 0.92 (0.90, 0.94) |
| **BMI** | **< 25** | 30 | 0.97 (0.96, 0.98) | 0.70 (0.65, 0.75) | 0.83 (0.77, 0.90) | 0.93 (0.91, 0.95) |
| | **≥ 25** | 11 | 0.97 (0.96, 0.98) | 0.68 (0.61, 0.75) | 0.81 (0.75, 0.88) | 0.94 (0.92, 0.96) |
| **Skin Tone** | **I, II, III** | 21 | 0.98 (0.97, 0.98) | 0.69 (0.65, 0.72) | 0.86 (0.82, 0.89) | 0.93 (0.92, 0.95) |
| | **IV, V, VI** | 20 | 0.94 (0.91, 0.98) | 0.73 (0.62, 0.92) | 0.75 (0.63, 0.90) | 0.93 (0.89, 0.98) |
| **Arm Hair Index** | **1** | 17 | 0.96 (0.94, 0.98) | 0.69 (0.63, 0.75) | 0.78 (0.68, 0.88) | 0.94 (0.93, 0.96) |
| | **2** | 16 | 0.97 (0.96, 0.98) | 0.73 (0.66, 0.82) | 0.85 (0.79, 0.90) | 0.94 (0.92, 0.97) |
| | **3, 4** | 8[a] | NA | NA | NA | NA |

[a]Due to insufficient number of samples (8) for this subgroup, we did not evaluate the performance
BMI = body mass index; CI= confidence interval; NPV = negative predictive value; PPV = positive predictive value

Supplementary Table 5. Performance of 4-class sleep stage classification for participant subgroups (BMI: body mass index; CI: confidence interval).

| Subgroup (n) | | n | Overall Cohen's Kappa (95% CI) | Light Sleep Kappa (95% CI) | Deep Sleep Kappa (95% CI) | REM Kappa (95% CI) | Wake Kappa (95% CI) |
|---|---|---|---|---|---|---|---|
| **Age** | **18-40 yrs** | 25 | 0.70 (0.69, 0.71) | 0.63 (0.42, 0.78) | 0.70 (0.37, 0.90) | 0.74 (0.53, 0.87) | 0.72 (0.50, 0.90) |
| | **> 40 yrs** | 16 | 0.63 (0.62, 0.64) | 0.56 (0.24, 0.73) | 0.60 (0.12, 0.9) | 0.73 (0.33, 0.9) | 0.66 (0.40, 0.85) |
| **Sex** | **Female** | 23 | 0.66 (0.65, 0.67) | 0.59 (0.26, 0.75) | 0.66 (0.23, 0.87) | 0.73 (0.35, 0.90) | 0.70 (0.41, 0.89) |
| | **Male** | 18 | 0.68 (0.67, 0.69) | 0.61 (0.42, 0.78) | 0.66 (0.26, 0.92) | 0.74 (0.53, 0.89) | 0.69 (0.50, 0.88) |
| **BMI** | **< 25** | 30 | 0.67 (0.67, 0.68) | 0.6 (0.27, 0.78) | 0.67 (0.24, 0.91) | 0.73 (0.34, 0.90) | 0.70 (0.42, 0.90) |
| | **≥ 25** | 11 | 0.67 (0.65, 0.68) | 0.59 (0.43, 0.73) | 0.63 (0.26, 0.87) | 0.74 (0.65, 0.84) | 0.68 (0.53, 0.82) |
| **Skin Tone** | **I, II, III** | 21 | 0.70 (0.69, 0.71) | 0.62 (0.45, 0.78) | 0.67 (0.28, 0.92) | 0.77 (0.61, 0.90) | 0.71 (0.52, 0.85) |
| | **IV, V, VI** | 20 | 0.64 (0.63, 0.65) | 0.57 (0.24, 0.74) | 0.65 (0.20, 0.87) | 0.70 (0.30, 0.88) | 0.68 (0.40, 0.90) |
| **Arm Hair Index** | **1** | 17 | 0.65 (0.64, 0.66) | 0.59 (0.28, 0.78) | 0.62 (0.19, 0.88) | 0.74 (0.38, 0.89) | 0.69 (0.41, 0.84) |
| | **2** | 16 | 0.689 (0.67, 0.69) | 0.60 (0.37, 0.75) | 0.69 (0.33, 0.87) | 0.73 (0.49, 0.89) | 0.72 (0.51, 0.91) |
| | **3,4** | 8[a] | NA | NA | NA | NA | NA |

[a]Due to insufficient number of samples (8) for this subgroup, we did not evaluate the performance
BMI = body mass index; CI= confidence interval
